# Localizing epileptogenic zones using interictal intracranial electroencephalography and deep learning

**DOI:** 10.64898/2026.07.22.26358705

**Authors:** Zekai Qiang, Francois Okoroafor, Rory J. Piper, Damjan Velanoski, Friederikke Moeller, Gerald Cooray, Krishna Das, Christin Eltze, Suresh Pujar, M Zubair Tahir, Richard Rosch, Martin Tisdall, Aswin Chari

## Abstract

**Introduction:** Approximately 25% of the 51.7 million people with epilepsy globally develop drug-resistant disease, for whom surgical resection offers a potential path to seizure freedom contingent on accurate localization of the epileptogenic zone (EZ). Current practice relies on ictal intracranial EEG (iEEG) monitoring, yet the extend of the seizure onset zone, delineated in this way does not reliably predict surgical outcomes. Most existing machine learning approaches exploit single-channel features involved in ictal onset, which fail to capture the network-level topology of the interictal period. None have demonstrated generalization across implantation modalities or clinical centers.

**Methods:** We developed a deep learning architecture operating on multichannel interictal iEEG, comprising a Morlet wavelet temporal Transformer encoder and a permutation-equivariant Induced Set Attention Block spatial encoder modelling long-range inter-electrode interactions. The model was evaluated on 50.5 hours of iEEG from 161 patients across 17,012 channels at seven independent centers, using leave-one-center-out (LOCO) cross-validation. The EZ was defined by the overlap between the clinical seizure onset zone and the resected area in patients achieving Engel Class I post-surgical outcome. Performance was benchmarked against a literature-derived pooled AUROC from a systematic review and meta-analysis of 11 published studies (46 study arms), and against electrophysiological event-rate baselines including IED rate and IED-HFO co-occurrence rate.

**Results:** The model achieved a pooled AUROC of 0.778 (95% CI: 0.748–0.808), comparable to both the literature benchmark (0.765; 95% CI: 0.743–0.787) and the IED rate baseline (0.782; 95% CI: 0.750–0.814), with above-chance discrimination at all seven held-out centers (per-center AUROC: 0.668–0.925). Classical machine learning classifiers applied to spectral features under LOCO cross-validation performed substantially below both the electrophysiological baselines and the literature benchmark, with the best-performing classifier (XGBoost) achieving an AUROC of 0.642. Implantation modality (SEEG vs. ECOG; p = 0.494), vigilance state (sleep vs. wakefulness; p = 0.350), and age group (pediatric vs. adult; p = 0.202) did not significantly affect model performance. A greater proportion of channels designated as EZ was the only patient-level variable inversely associated with performance (ρ = −0.192, p = 0.015). Extraction of model attention scores allowed interrogation of discriminative interictal epochs.

**Discussion:** Model performance was comparable to established electrophysiological baselines and the literature benchmark under cross-center generalization, with performance variability attributable to principally EZ spatial extent. Model attributions aligned with established interictal electrophysiology. Limitations include the retrospective design, a predominantly pediatric cohort, and the absence of structural neuroimaging or effective connectivity priors.

**Conclusion:** A temporally and spatially aware deep learning architecture can localize the EZ from interictal iEEG with consistent cross-center and cross-modality generalization, performing comparably to established electrophysiological biomarkers without requiring manual annotation. These findings establish a foundation for AI-assisted EZ hypothesis generation and motivate prospective validation in clinical presurgical workflows.

## Introduction

Epilepsy is a chronic neurological disorder affecting approximately 51.7 million people globally (1). Although antiepileptic medications achieve adequate seizure control in the majority of patients, approximately 25% develop drug-resistant epilepsy (DRE) (2). In a carefully selected subset of patients with DRE, surgical resection offers a potential route to seizure freedom, contingent upon accurate localization of the epileptogenic zone (EZ). The EZ is defined as the area of cortex that is both necessary and sufficient for the initiation of seizures, and whose removal or disconnection is required for their complete abolition (3). As the EZ can only be definitively confirmed following successful surgery that renders a patient seizure-free, the seizure onset zone (SOZ), which is defined as the brain region from which clinical seizures are first electrographically observed, serves as its primary clinical surrogate (4). In selected candidates, the SOZ is identified through subacutely implanted intracranial electroencephalography (iEEG) using stereoelectroencephalography (SEEG) or subdural electrocorticography (ECOG) grids and strips, wherein clinical neurophysiologists review electrographic patterns to identify the earliest ictal changes (5). It is important to recognize that the implantation strategy itself introduces a sampling bias and limitation to this concept of SOZ, because only regions within or proximate to implanted electrode trajectories can be assessed, and cortex outside the implant footprint remains electrographically invisible. Therefore, the SOZ alone may not accurately delineate the full spatial extent of the EZ, and the proportion of SOZ contacts included within the surgical resection does not reliably correlate with post-operative seizure freedom (6, 7).

This discrepance between SOZ-based surgical planning and clinical outcomes reflects a fundamental conceptual limitation, as epilepsy is increasingly understood as a network disorder characterized by an underlying predisposition to generate seizures (8). The delineation of the irritative zone, representing the cortical region generating interictal epileptiform discharges (IEDs), forms a component of pre-surgical evaluation (9). Indeed, IEDs may themselves be regarded as electrographic biomarkers of epileptogenicity reflecting the network’s inherent susceptibility to seizure generation (10). Nevertheless, current approaches to invasive monitoring remain principally oriented towards the capture of spontaneous ictal events, and there is potential to operationalize interictal recordings to provide a more comprehensive characterization of epileptogenic tissue. Advanced analytical methods that can extract robust biomarkers of epileptogenicity from interictal iEEG data therefore may localize EZ beyond traditional ictal-based approaches. In time, reliable interictal localization may also reduce dependence on prolonged ictal recording and length of hospital stay.

Machine learning (ML) approaches have been increasingly applied to the characterization of the SOZ using quantitative representations of iEEG data. For a comprehensive review of the current ML approaches adopted and the current performance benchmark, see Supplemental Material 2. Statistical, feature-based ML methods exploit a predefined repertoire of biomarkers computed from iEEG recordings, including rates of high-frequency oscillations (HFOs) and IEDs (11-15), spectral power estimates (16-19), entropy-derived measures (20, 21), and graph-theoretic network features such as degree centrality, clustering coefficient, and local efficiency (16, 22-24). Reported performance across these studies varies considerably, with area under the receiver operating characteristic curve (AUROC) values generally ranging from approximately 0.70 to 0.85 (Supplemental Material 2). Deep learning (DL) approaches, which discover classification boundaries in large datasets through backpropagation-guided hierarchical representation learning, (25) have similarly been explored. Current efforts included convolutional neural networks with data augmentation (26), adversarial neural networks with domain adaptation (27), graph neural networks (28), and semi-supervised convolutional autoencoders (29). However, published DL models have not consistently demonstrated performance superiority over well-engineered feature-based classifiers and are difficult to contextualize given the absence of standardized benchmarking. No existing study has rigorously evaluated the generalization of DL architectures across implantation modalities or recording centers.

In this study, we propose a temporally and spatially aware DL architecture for the characterization of the EZ from interictal iEEG recordings, and evaluate its performance in a multi-centric, offline setting. We also benchmark it against established clinical neurophysiological methods, classical ML methods, and the current literature reported performances.

## Results

### Dataset characteristics

A total of 50.5 hours of interictal iEEG are analyzed across 161 subjects and 17,012 channels, from seven clinical centers: GOSH (n=14), NIH (n=8), JHU (n=3), UMF (n=3), UPenn (n=33), Detroit (n=86), and UCLA (n=14). (Figure 1, Supplemental Material 4) The cohort comprised 47 adults and 114 pediatric patients, with more recordings obtained during sleep (n=100) than wakefulness (n=61). Implantation devices varied by center, with 121 subjects implanted with ECOG grids and 40 with SEEG depth electrodes; GOSH and Detroit employed exclusively one modality (SEEG and ECOG, respectively), whereas NIH, JHU, and UPenn utilized both. Available recording durations differed across centers, ranging from a mean of 2.03 minutes (UMF) to 89.07 minutes (UCLA). The proportion of channels designated as the EZ also varied across sites, with a mean of 9.07% (SD=10.04%) across all subjects; UMF exhibited a higher EZ channel proportion (mean 53.07%, SD=11.50%) compared to all other centers, where means ranged from 6.38% to 17.57%.

**Figure 1.**
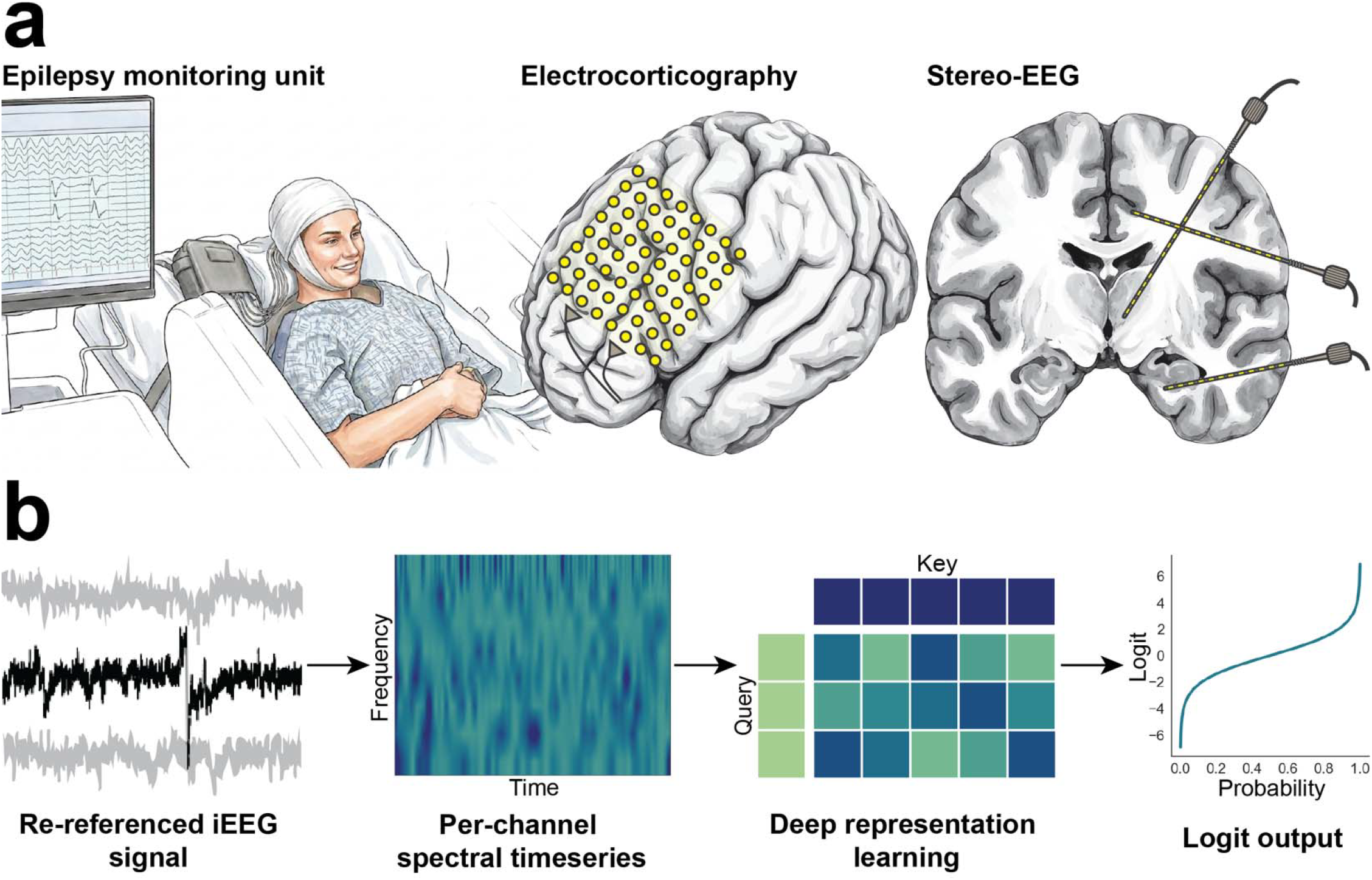
Intracranial electroencephalography (iEEG) data flow. (a) Interictal iEEG data is acquired from patients undergoing pre-surgical evaluation for epilepsy and can be obtained through electrocorticography (ECOG) or stereo-electroencephalography (SEEG). (b) The recorded SEEG is re-referenced by taking the bipolar montage and ECOG by subtracting common mean across the electrode grid. Temporally resolved spectral features are calculated for each channel forming the tensor subjected to the deep learning (DL) framework. The DL framework operates on the transformer backbone, and a schema of the self-attention mechanism is shown. The DL pipeline returns per-channel logit output representing their likelihood of being part of the epileptogenic zone.

**Figure 2.**
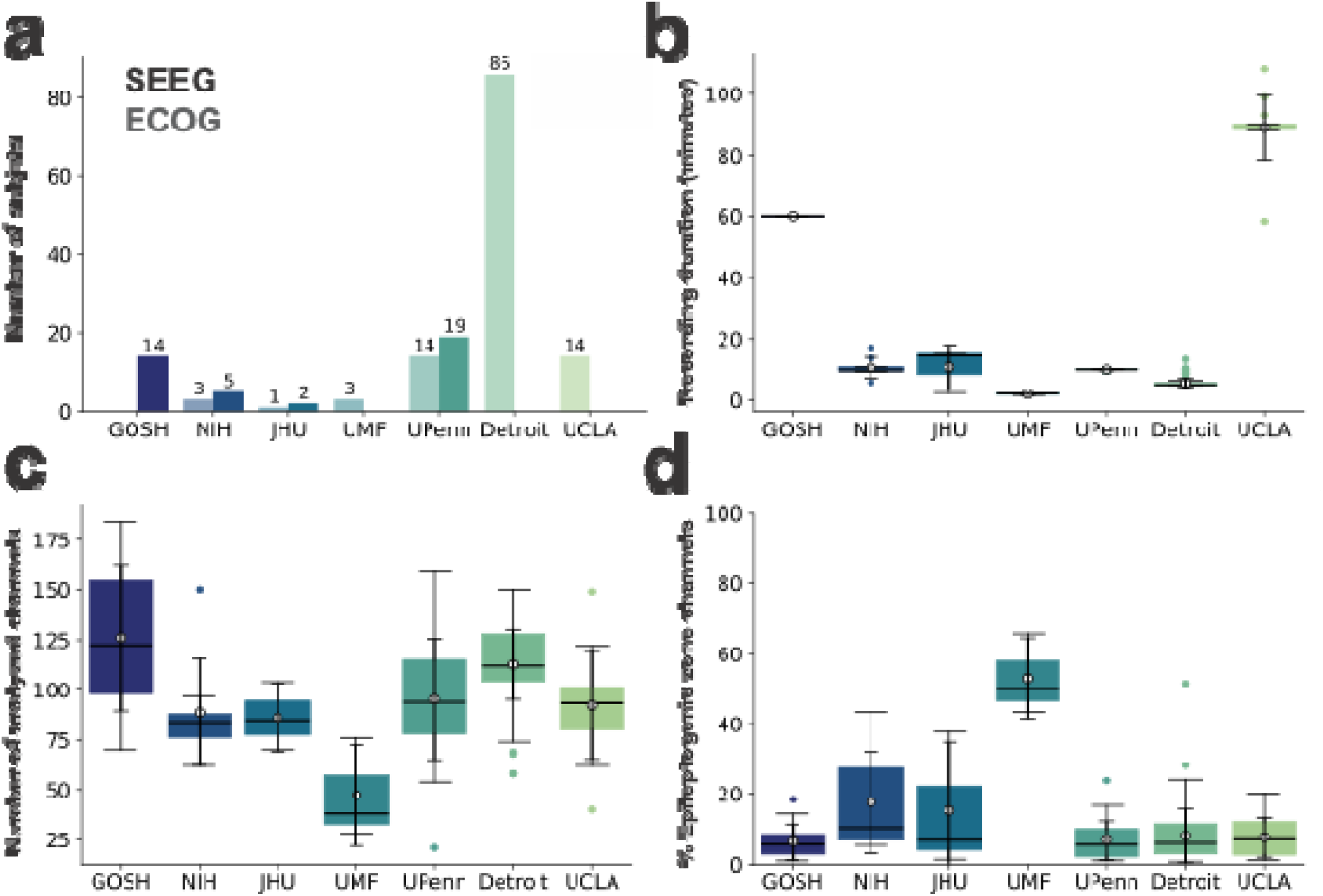
Dataset characteristics. (a) A total of 50.5 hours of interictal intracranial EEG were analyzed across 161 subjects (17,012 channels) from seven clinical centers. Implant type varied by centers. 121 subjects received ECOG grids and 40 received SEEG depth electrodes. The darker bars to the right of the tick labels show the number of patients implanted with SEEG, and the paler colors to the left of the tick labels show the number of ECOG patients. (b) Recording duration differed substantially across centers, ranging from a mean of 2.03 min (UMF) to 89.07 min (UCLA). (c) The number of analyzed channels in each subject per center ranged from 21 to 184. (d) The proportion of channels designated as the epileptogenic zone (EZ) varied across sites, with a mean of 9.07% (SD = 10.04%) across all subjects.

**Figure 3.**
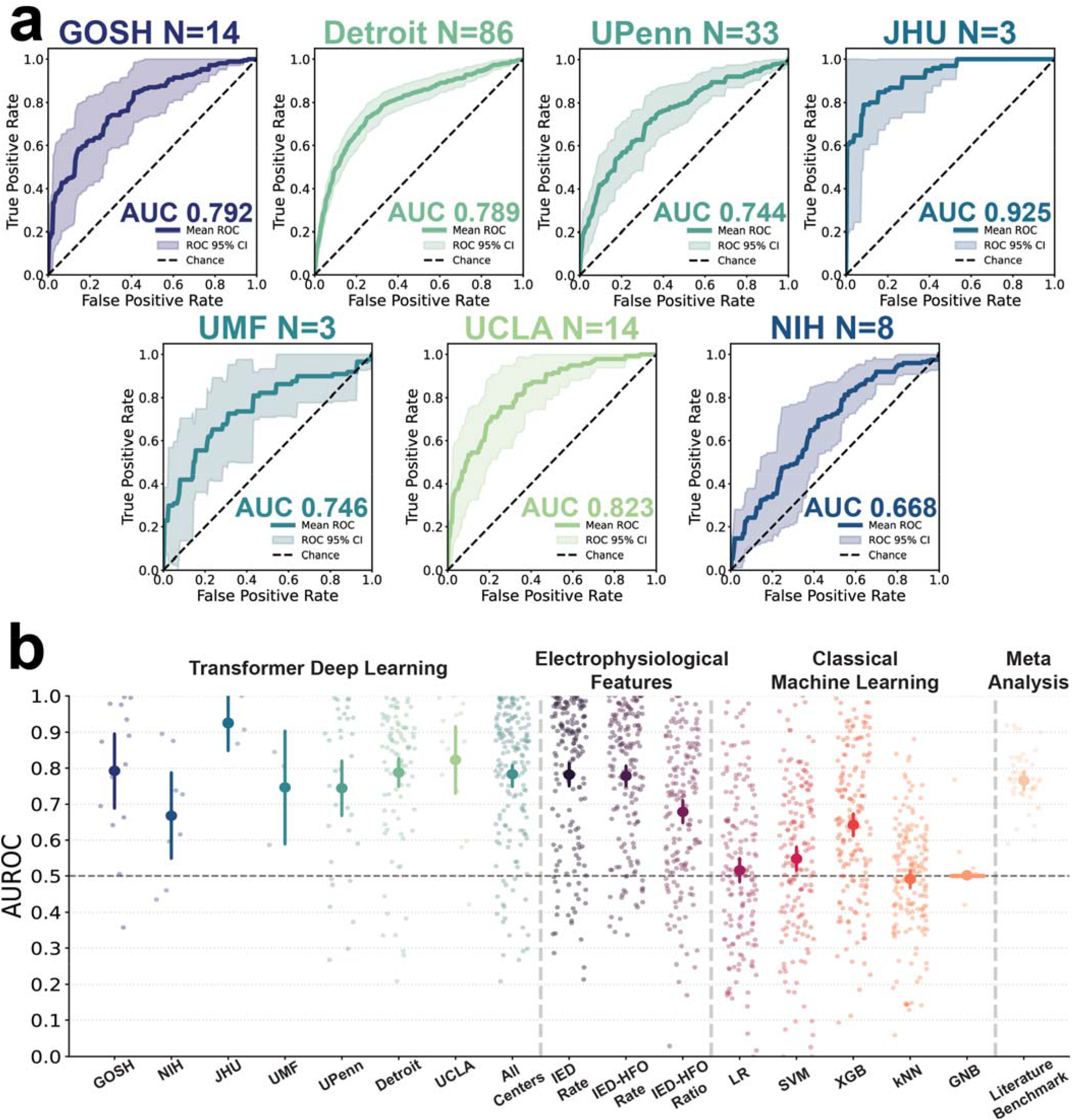
Performance of the deep learning model across centers and against benchmarks. (a) Receiver operating characteristic (ROC) curves demonstrating the performance of the proposed deep learning (DL) model under a leave-one-center-out validation framework. Model performance remained robust across all centers, achieving AUROC values of 0.792 (95% CI, 0.689–0.896) at GOSH (N = 14), 0.668 (95% CI, 0.549–0.787) at NIH (N = 8), 0.925 (95% CI, 0.848–1.000) at JHU (N = 3), 0.746 (95% CI, 0.588–0.904) at UMF (N = 3), 0.744 (95% CI, 0.668–0.820) at UPenn (N = 33), 0.788 (95% CI, 0.748–0.827) at Detroit (N = 86), and 0.823 (95% CI, 0.729–0.916) at UCLA (N = 14). (b) Comparison of the proposed DL model against established electrophysiological biomarkers, classical machine learning (ML) approaches, and a benchmark derived from a literature-based meta-analysis. Baseline electrophysiological features achieved AUROCs of 0.782 (95% CI, 0.750–0.814) for interictal epileptiform discharge (IED) rate, 0.778 (95% CI, 0.749–0.807) for IED-associated high-frequency oscillation (IED-HFO) rate, and 0.679 (95% CI, 0.648–0.710) for the IED-HFO ratio (N = 161). Classical ML models demonstrated substantially lower discriminative performance, including logistic regression (AUROC 0.516, 95% CI, 0.483–0.549), support vector machine (AUROC 0.548, 95% CI, 0.515–0.581), XGBoost (AUROC 0.642, 95% CI, 0.611–0.673), k-nearest neighbors (AUROC 0.492, 95% CI, 0.467–0.516), and Gaussian naïve Bayes (AUROC 0.502, 95% CI, 0.498–0.505) (all N = 161). The literature-derived meta-analytic benchmark yielded an AUROC of 0.765 (95% CI, 0.743–0.787; N = 46 study arms).

### Model performance across centers

The proposed DL model evaluated under the same LOCO cross-validation scheme, achieved a pooled AUROC of 0.778 (95% CI: 0.748–0.808; N = 161) across held-out subjects. Per-center AUROCs varied from 0.668 (95% CI: 0.549–0.787; N = 8) at the NIH to 0.925 (95% CI: 0.848–1.000; N = 3) at JHU, with intermediate performance observed at GOSH (0.792; 95% CI: 0.689–0.896; N = 14), UCLA (0.823; 95% CI: 0.729–0.916; N = 14), Detroit (0.788; 95% CI: 0.748–0.827; N = 86), UMF (0.746; 95% CI: 0.588–0.904; N = 3), and UPenn (0.744; 95% CI: 0.668–0.820; N = 33).

### Factors associated with model performance

Spearman rank correlation analysis revealed that neither the total number of input channels (ρ = −0.013, p = 0.872) nor the duration of the analyzed interictal recording (ρ = 0.076, p = 0.337) significantly predicted individual patient-level model performance. Recordings obtained during sleep were not associated with significantly different performance compared with those obtained during wakefulness (Mann–Whitney U = 3318.50, p = 0.350), and no significant difference in performance was observed between SEEG and ECOG implantation modalities (U = 2244.50, p = 0.494), nor between adult and pediatric cohorts (U = 2335.50, p = 0.202). However, the proportion of channels classified as within the EZ was inversely associated with AUROC (ρ = −0.192, p = 0.015).

### Interpretability analysis

Interpretability analysis revealed that the model identifies patient-level, discriminative waveform morphologies that parallel established interictal electrographic patterns (Figure 4). Several high-attention clusters exhibited a sharp, high-amplitude deflection followed by a broader, lower-frequency slow wave, potentially consistent with the classic spike-and-slow-wave morphology of IEDs. Others showed repetitive, semi-regular oscillatory bursts of moderate amplitude, resembling rhythmic paroxysmal discharges. These findings support the interpretability and clinical utility of the model, suggesting that its predictions can be traced to recognizable electrographic patterns and may therefore offer clinicians a transparent, waveform-level rationale to complement EZ localization decisions.

**Figure 4.**
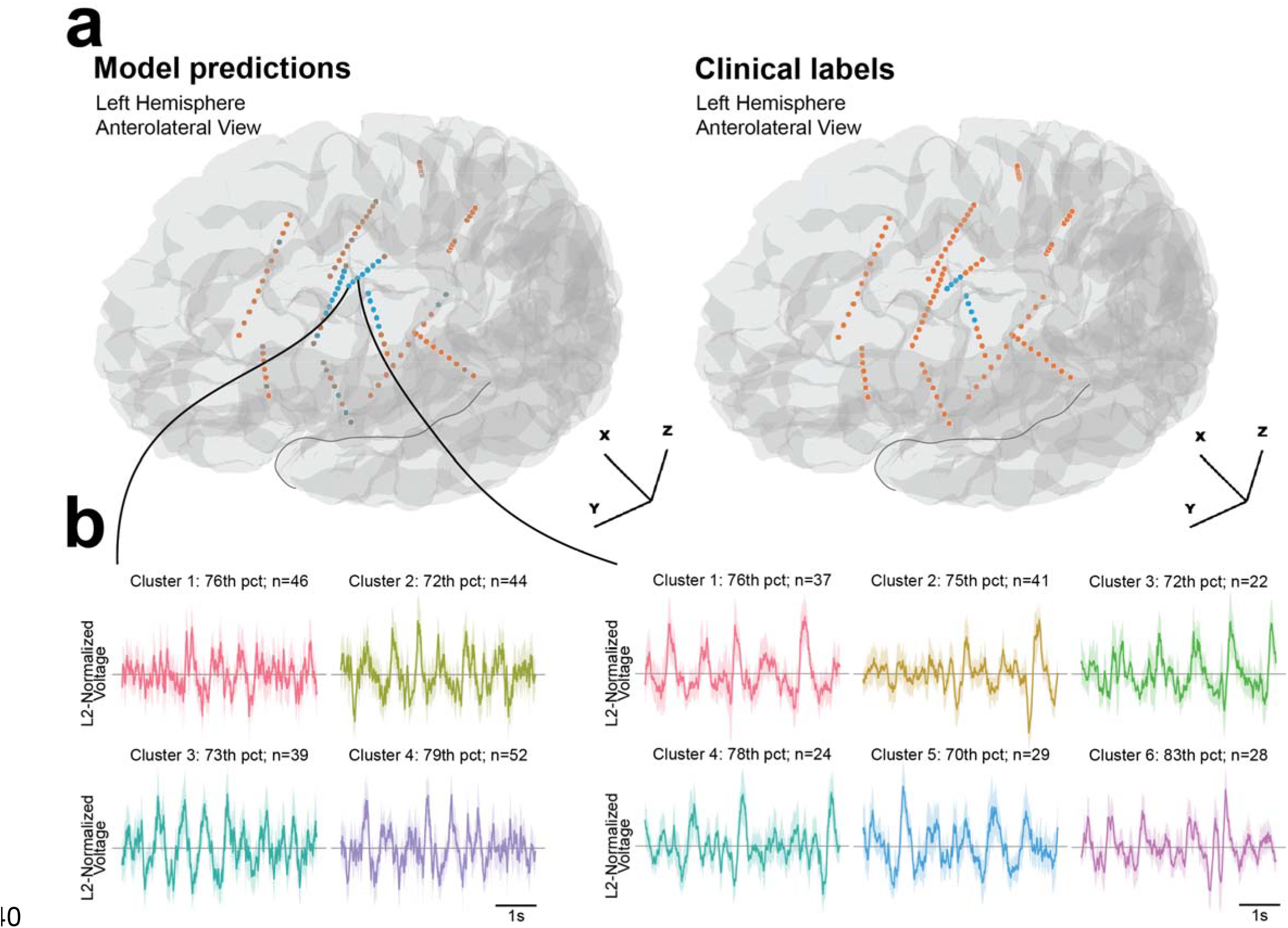
Interpreting deep learning based epileptogenic zone classification. (a) iEEG bipolar channels for a representative patient from the GOSH cohort. Left panel demonstrates model-predicted epileptogenic zone (EZ) channels (blue) versus non-EZ channels (orange). Right panel demonstrates corresponding clinician-assigned ground-truth labels for the same electrode array. (b) Cluster-derived discriminative waveform prototypes from the channel-level interpretability pipeline, shown for two example channels. Each panel shows the mean L2-normalized voltage trace (± 95% CI) for one morphological cluster identified via unsupervised agglomerative clustering of high-attention epochs, labeled by the percentile rank of the cluster’s median attention weight relative to the full attention distribution for that channel (pct) and the number of constituent epochs (n). Four clusters derived from epochs selected using model-predicted attention were shown for one example EZ channel, compared to six clusters identified in a different EZ channel. Similar recurring waveform morphologies across both sets of clusters indicate that the model attends to physiologically plausible, clinically concordant epileptiform patterns rather than spurious or artifactual features.

### Establishing baseline comparators

To compare the performance of the proposed DL architecture within the broader landscape of published approaches, a systematic review and random-effects meta-analysis was conducted encompassing 11 studies (46 study arms). The pooled AUROC derived from the literature was 0.7650 (95% CI: 0.7384–0.7897). (Supplemental Material 2)

Electrophysiological event-rate features demonstrated competitive discriminative performance. The IED rate achieved an AUROC of 0.782 (95% CI: 0.750–0.814; N = 161), the rate of IEDs co-occurring with high-frequency oscillations (IED-HFO rate) yielded a closely comparable AUROC of 0.778 (95% CI: 0.749–0.807). The IED-HFO ratio was the least discriminative of the three electrophysiological features, achieving an AUROC of 0.679 (95% CI: 0.648–0.710).

Classical ML classifiers applied to spectral features under LOCO cross-validation performed substantially below both the electrophysiological baselines and the literature benchmark. Gradient-boosted trees were the strongest-performing classical classifier at an AUROC of 0.642 (95% CI: 0.611–0.673), followed by the support vector machine with radial basis function kernel at 0.548 (95% CI: 0.515–0.581). Logistic regression, k-nearest neighbors, and Gaussian naïve Bayes all performed near chance level, achieving AUROCs of 0.516 (95% CI: 0.483–0.549), 0.492 (95% CI: 0.467–0.516), and 0.502 (95% CI: 0.498–0.505), respectively.

## Discussion

We present a hierarchical DL architecture integrating channel-level temporal encoding with permutation-equivariant spatial reasoning for EZ localization from interictal iEEG. Evaluated under LOCO cross-validation across 161 subjects from seven individual centers, the model achieved a pooled AUROC of 0.778 (95% CI: 0.748–0.808). This was comparable to a literature-derived benchmark of 0.765 (95% CI: 0.743–0.787), established via systematic review and random-effects meta-analysis of 46 study arms from 11 published studies, and to the IED rate baseline of 0.782 (95% CI: 0.750–0.814) computed on the same cohort.

Per-center AUROC ranged from 0.668 to 0.925, confounded by heterogeneity in electrode implantation strategy, patient demographics, recording duration, and class imbalance. Despite this variability, above-chance discrimination was achieved at every center. Spearman correlation analysis identified the proportion of channels classified as epileptogenic as the only patient-level variable inversely associated with performance (ρ = −0.192, p = 0.015), suggesting that diffuse or spatially distributed epileptogenic networks may present a harder classification problem than well-circumscribed focal zones.

### Spatial architecture

Prior DL approaches to putative EZ localization have mostly operated on single-channel representations which neglects the spatial topology of epileptic networks. Several localizing networks have been identified in the literature. For instance, SOZ regions demonstrate significantly greater inward strength, potentially representing interictal suppression, with notably weaker information flow from SOZ to non-SOZ across a broad frequency range (1–250 Hz) (30, 31). Such spatial topology may even carry prognostic information, as greater within-frequency information flow asymmetry between SOZ and non-SOZ were found associated with more favorable seizure outcomes (30). In contrast, effective connectivity studies using single-pulse electrical stimulations applied to EZ have demonstrated strongest responses both within and outside these zones, characterizing its high excitability and dominant influence over other regions within the epileptogenic network (32). Other investigations found that the SOZ is densely self-connected with minimal input from non-SOZ regions, suggesting relative isolation from surrounding cortex (33, 34). The present architecture addresses this gap through a spatial encoder that is permutation-equivariant, making no assumptions about electrode ordering, anatomical labelling, or implant geometry.

### Literature-derived benchmark

Of the 24 studies identified in our systematic review, 11 reported AUROC estimates, yielding 46 independent study arms. The resulting pooled AUROC of 0.765 (95% CI: 0.743–0.787) constitutes the first formally derived literature benchmark for this task. Stratified analysis by algorithm class yielded pooled AUROCs of 0.758 (95% CI: 0.731–0.782) for classical ML (k = 42 arms) and 0.810 (95% CI: 0.694–0.889) for DL (k = 4 arms). While the DL point estimate is numerically superior, confidence intervals overlapped substantially and the DL interval was considerably wider, reflecting the scarcity of available study arms.

Moderate between-study heterogeneity was detected (I^2^ ≈ 0.33), attributable principally to inconsistent positive class definitions. Across reviewed studies, the positive class variously encompassed the clinician-determined SOZ regardless of outcome, the intersection of resected and seizure onset volumes, and the resected area in seizure-free patients. Each of these represent a conceptually and operationally distinct labeling construct and are limited by the iEEG implantation bias a priori. Where positive labels include electrodes from patients with surgical failure, labeled contacts may not be causally necessary for seizure generation, attenuating observed performance. Additional sources of heterogeneity included wide variation in sample size (3–101 patients), recording modality, vigilance state, epilepsy etiology, class imbalance, and feature engineering. These considerations suggest the pooled benchmark is best interpreted as a population-level central tendency rather than a precise performance target and motivated the outcome-validated EZ definition adopted in the present study.

### Baseline comparators

Both electrophysiological event-rate features and classical ML classifiers applied to Morlet-derived spectral power features were evaluated on the same cohort under identical LOCO cross-validation. Among electrophysiological baselines, IED rate was the strongest individual feature (AUROC 0.782, 95% CI: 0.750–0.814), marginally exceeding the pooled literature estimate. IED-HFO co-occurrence rate performed comparably (0.778, 95% CI: 0.749–0.807), while the IED-HFO ratio was least discriminative (0.679, 95% CI: 0.648–0.710).

The competitive performance of a simple, computationally parsimonious IED rate against the full DL architecture warrants reflection. It suggests that a substantial portion of the discriminative signal accessible from interictal iEEG may already be captured by this single interpretable measure. This however should not diminish the DL approach, whose architecture is in principle capable of capturing temporal dynamics, cross-band interactions, and cross-channel spatial relationships inaccessible to univariate rate summarization. Although it does imply that advances in model performance may require richer signal representations, improved EZ ground truth definitions, or integration of complementary data sources. Practically, the DL model’s ability to operate on raw multichannel recordings without manual IED annotation remains a clinically relevant advantage, reducing the labor and inter-rater variability of clinical neurophysiological review.

### Limitations

Several limitations merit consideration. Evaluation was performed in an offline, retrospective setting. Prospective validation within live clinical localization workflows is essential. The LOCO framework simulates deployment to a new center but does not account for distribution shift arising from hardware upgrades, evolving clinical practice, or novel implantation strategies over time. The EZ label is bounded by the extent of clinical implantation, as channels outside the implanted array remain invisible to the model, and this is a fundamental constraint of any iEEG-based approach. The current architecture does not incorporate structural connectivity, neuroimaging, effective connectivity via single-pulse electrical stimulation, or electrode coordinates. Integrating MRI-derived parcellations, brain stimulation evoked potentials, or MNI coordinates as inductive priors could further constrain the model’s attention and improve performance.

## Conclusions

Taken together, this study provides a comprehensive and rigorously benchmarked evaluation of interictal DL for EZ localization, achieving robust generalization across institutions and patient populations, and establishing both a standardized cross-center evaluation framework and the first formally derived literature benchmark for this task. These contributions lay the groundwork for future validation and the integration of data-driven interictal biomarkers into the presurgical evaluation of drug-resistant epilepsy.

## Materials and Methods

### iEEG acquisition

iEEG recordings are obtained from 161 patients with drug-resistant epilepsy across seven tertiary centers: Great Ormond Street Hospital for Children (GOSH), Johns Hopkins University (JHU), the University of Pennsylvania (UPenn), U.S. National Institutes of Health (NIH), University of Miami Florida Jackson Memorial Hospital (UMF), University of California Los Angeles Mattel Children’s Hospital (UCLA), and Children’s Hospital of Michigan (Detroit). (6, 35-37)

### iEEG preprocessing

Raw iEEG recordings are first high pass filtered to remove slow drifts, followed by notch filtering at either 50 Hz (GOSH cohort) or 60 Hz (NIH, JHU, UPenn, UMF, UCLA, Detroit cohorts) according to the mains frequency of the recording institution to attenuate power line interference. Bipolar re-referencing is applied to SEEG recordings by computing differential voltages between adjacent contacts along each electrode shaft. Common average re-referencing is applied to ECOG recordings wherein the mean signal across all electrodes on the same grid was subtracted from each individual channel. Signals are then downsampled using zero-phase decimation with anti-aliasing filtering to achieve the target sampling rate of 256 Hz.

### Epileptogenic zone definition

In the present study, the EZ is defined as channels meeting two criteria: (1) identification as part of the SOZ by clinical neurophysiologists, and (2) subsequent resection or ablation in patients who achieved Engel Class I seizure freedom outcomes. This definition ensured that only causally validated epileptogenic tissue was labeled as positive examples for DL. For each bipolar pair, channel labels are propagated such that a bipolar channel was labeled as EZ if either constituent contact was part of the EZ. This labelling strategy ensured that all potentially epileptogenic tissue is captured in the positive class.

### Model Architecture Overview

We developed a hierarchical DL architecture that operates directly on pre-processed, multichannel time-series signals and produces a per-channel scalar logit reflecting the likelihood of EZ candidacy. The architecture comprises three sequentially composed modules: (i) a temporal encoder, which transforms each channel’s raw signal into a fixed-dimensional embedding via a bank of complex Morlet wavelets followed by a hierarchical Transformer (38), (ii) spatial transformer layers, which model long-range interactions across the full electrode array in a permutation-equivariant manner, and (iii) a per-channel classification head, which maps each temporally and spatially integrated channel embedding to a scalar EZ logit. The model hyperparameters are subject to configuration, and those used in the currently reported inference run is detailed in Supplemental Material 3.

### Temporal encoding

Each channel’s raw iEEG signal is first decomposed into a multi-band time–frequency representation using a bank of complex Morlet wavelets with center frequencies distributed logarithmically from 1 to 120 Hz. Six wavelet bands are employed, each with an initial cycle count of 5. The real and imaginary parts of each wavelet are energy-normalized and convolved with the pre-processed signal, and the instantaneous magnitude per band is computed.

To mitigate cross-subject and cross-electrode confounds in raw wavelet magnitude, a band-wise log-power standardization is applied within each channel. For each band, the instantaneous log-power series is computed and subsequently z-scored across the full recording duration. The standardized scores are then symmetrically clipped to [-20, 20] for numerical stability during training, and passed through the shifted exponential non-linearity *f(z)= e*^*Z*^ *-1*, which re-expands dynamic range while preserving rank ordering of band activity.

To adapt to the variable recording durations available in the dataset, the temporal encoder adopts a two-stage hierarchical architecture that segments the recording into non-overlapping epochs of fixed duration (10 seconds), encodes each epoch independently, and then aggregates across epochs. Each epoch tensor is adaptively average-pooled along the time axis to *K* = 8 temporal tokens and are projected into the model dimension *d* = 64. Standard sinusoidal positional encodings are added, followed by a single-layer Transformer encoder to apply self-attention across temporal tokens. The epoch-level embedding is then obtained by max-pooling the token sequence along the temporal dimension. Across-epoch attention aggregation is applied to transform length-dependent token sizes into single, fixed-size vector using a single learnable query vector.

### Spatial encoding

The spatial module must capture pairwise relationships between electrode channels without assumptions about spatial proximity, channel ordering, or anatomical labelling. The Induced Set Attention Block (ISAB) (39) of the Set Transformer framework is adopted to satisfy this permutation-equivariance requirement whilst avoiding the *O*(*C*^2^) complexity of naive all-to-all attention.

Given the channel temporal embedding matrix, the ISAB introduces 32 learnable inducing points. The computation proceeds in two attention steps via a multihead attention block. In the first block, the inducing points serve as queries and the channel embeddings as keys and values, compressing the full channel set into a compact latent representation. In the second block, the channel embeddings serve as queries against the updated inducing points, expanding the compressed representation back to the full channel set. Both blocks employ 2 attention heads, with each head operating on a dimension of 32.

### Feedforward layer and classification

The ISAB output is processed by a position-wise feed-forward network employing the SwiGLU activation function. A task-specific classification head independently maps each channel’s contextualized embedding to a scalar EZ logit. The head is implemented as a two-layer multi-layer perceptron with intermediate dimension 32, a second dropout layer (*p* = 0.1) applied before the final linear projection, and a bias term on the output layer. The vector of channel-wise logits is passed through a sigmoid function to produce per-channel likelihood at inference time.

### Composite loss function

Training optimizes a composite objective combining a primary per-channel binary classification term with an auxiliary pairwise ranking term:

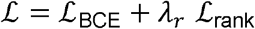

where *λ*_*r*_ = 0.2 is the ranking loss weight.

### Leave-one-center-out cross validation

To assess generalization to unseen clinical centers, a LOCO cross-validation scheme was employed. In each fold, all subjects from a single recording center were withheld as the held-out evaluation set, and the model was trained from scratch on subjects from all remaining centers. This procedure was repeated until every center had served exactly once as the held-out site.

Class-imbalance weights are recomputed independently for each training fold based solely on the label distribution of subjects in the current training partition, ensuring that no label statistics from the held-out center entered the loss function.

Model performance was evaluated at the per-channel level using the AUROC, computed independently for each subject in the held-out center.

### Interpretability analysis

To evaluate whether the model relies on physiologically plausible iEEG features, a channel-level interpretability analysis was performed in which per-epoch attention weights were extracted from the trained transformer’s temporal encoder for each 10-second epoch of a given channel’s recording, yielding a time-resolved attention profile across the full recording. Epochs falling at or above the 50th percentile of attention were selected as high-salience segments, and their raw waveforms were extracted and L2-normalized to remove amplitude scaling effects prior to morphological comparison. Discriminative waveform morphologies were then identified in a fully unsupervised manner using agglomerative hierarchical clustering (average linkage) on a pairwise cosine-distance matrix, with the optimal number of clusters (k = 2–8, minimum cluster size ≥10% of segments) selected via the silhouette score. This allows assessment for whether high-attention epochs corresponded to distinct, recurring waveform morphologies consistent with known physiological signatures.

### Establishment of baseline comparators

To contextualize the performance of the proposed DL architecture, a set of baseline comparators is established using clinical neurophysiological features, classical ML with statistical summary of wavelet timeseries, and AUROC values reported in the current literature. For methodological details on how IED, HFO, IED-HFO coupling features are computed, and evaluation of classical ML methods, see Supplemental Material 1. For methods in deriving a literature-based benchmark of existing ML models, see Supplemental Material 2.

## Supporting information

Supplementary Material 1: Deriving electrophysiological and classical machine learning baseline comparators

Supplementary Material 2: Developing a benchmark for machine learning based localization of putative epileptogenic zones from human interictal intracr

Supplementary Material 3: Model hyperparameters

Supplementary Material 4: Cohort Clinical Information

## Data Availability

All data produced in the present study are available upon reasonable request to the authors.

## Author contributions

