## Supplementary Material 1: Deriving electrophysiological and classical machine learning baseline comparators for "Localizing epileptogenic zones using interictal intracranial electroencephalography and deep learning"

To benchmark the presented deep learning model against interpretable, clinically established features, we extract two feature sets: an electrophysiological set producing interictal event-rate features, and a spectral set producing Morlet-derived band-power features. The spectral feature set is then input to classical machine learning classifiers under a leave-one-center-out (LOCO) cross-validation scheme, matching that used for the Transformer.

Interictal Epileptiform Discharge (IED) Detection

The IED detector implements Janca et al. 2015 (1) which discriminates salient discharges from background activity using an adaptive modelling of signal envelop distribution. Methodological details can be found at (1) and original code at <https://github.com/EpiReC-ISARG/IED_detector>. The detection procedure and the parameters used to derive per-channel IED rate in the current study is summarized here:

1. The iEEG signal is zero-phase band-pass filtered using a 4th-order Butterworth filter with a passband of 10–70 Hz to eliminate phase distortion. This attenuates slow background drift and high-frequency noise while preserving the morphological characteristics of IEDs.
2. The Hilbert transform is applied to the filtered signal to obtain the analytic signal, and its instantaneous amplitude (envelope) is extracted.
3. The recording is partitioned into overlapping sliding windows of length 5 seconds, with a 1-second step. This yields a sequence of background-estimation epochs.
5. Within each window, the natural logarithm of the positive envelope samples is computed. The sample mean (μ) and sample standard deviation (σ) of the log-transformed envelope are estimated. The background amplitude distribution is modelled as lognormal.
6. The per-window μ and σ estimates are smoothed using a symmetric moving-average, then up-sampled to the full signal resolution by cubic spline interpolation. This produces continuous time-varying lognormal parameter traces μ(t) and σ(t).
7. Three time-varying lognormal statistics are derived from μ(t) and σ(t): mode: $exp(\mu-\sigma^{2})$, median: $exp(\mu)$, and mean: $exp(\mu+\sigma^{2}/2)$.
8. Adaptive amplitude threshold is constructed at each time point and is set as

$$\theta(t)=3\cdot(\text{mode}+\text{median})$$

9. For each threshold level, all contiguous segments where the envelope exceeds the threshold are identified. Within each segment, local envelope maxima are extracted by detecting sign changes in the first difference of the signal. For very short segments ($\leq2$ samples), the global maximum is selected instead.
10. Adjacent detected peaks separated by less than 0.12 seconds are merged via morphological dilation and erosion. This simulates the union of individual spikes within a polyspike complex. After merging, only the single highest-amplitude peak is retained per fused run.
11. The initial and terminal 1-second epochs (corresponding to filter ringing artefacts) are blanked from all marker arrays.
12. The IED rate per channel is expressed as event rate per minute.

High-Frequency Oscillation (HFO) Detection

HFOs are automatically detected within the band 80–120 Hz using a threshold-based procedure, following the Hilbert-envelope methodology described by Kucewicz et al. 2014 (2):

1. The single-channel signal is zero-phase band-pass filtered (4th-order Butterworth) within the HFO frequency band, by default 80–120 Hz
2. The instantaneous amplitude envelope is derived from the analytic signal of the band-passed signal.
3. A global, stationary amplitude threshold is computed as:

$$\theta_{\text{HFO}}=\mu_{\mathrm{Envelope}}+k_{\sigma}\cdot\sigma_{\mathrm{Envelope}}$$

where $\mu_{E}$ and $\sigma_{E}$ are the mean and standard deviation of the envelope across the entire recording, and $k_{\sigma}=3$ SD

4. Contiguous epochs during which $E(t)>\theta_{\text{HFO}}$ are identified as candidate HFO events.
5. Each candidate is subjected to two rejection criteria applied conjunctively:
   - The event must span at least = 10 milliseconds.
   - The event must contain at least 4 full oscillatory cycles at the lower band edge.

IED-HFO Coupling Detection

Coupling quantifies the temporal co-occurrence of HFOs with IED events.

1. For a given channel, IED peaks are obtained from the IED detector. HFO peaks are obtained from the HFO detector run independently on the same channel.
2. A coupling window of 50 milliseconds pre and post relative to each IED peak is defined.
3. For each IED peak, the set of HFO peaks falling within the coupling window is identified. If at least one HFO peak is present, the IED is classified as coupled.
4. Two scalar features are derived per channel:

IED–HFO rate: total coupled IEDs normalized by recording duration

$$\text{IED-HFO rate}=\frac{N_{\text{coupled}}}{T_{\text{min}}}$$

IED–HFO ratio: proportion of IEDs on that channel that were coupled to an HFO event

$$\text{IED-HFO ratio}=\frac{N_{\text{coupled}}}{max(N_{\text{IED}},\text{ }1)}$$

The denominator floor of 1 prevents division by zero for channels with no detected IEDs; such channels receive a ratio of 0.

5. Repeated independently for every channel in the recording, producing a per-channel vector of three electrophysiological features.

Classical Machine Learning Baseline

Spectral features were derived via analytic Morlet convolution analogous for input to the temporal encoding module. Two summary statistics are computed per channel, per band: the temporal mean and temporal standard deviation. This produces a feature vector of dimension 12 per channel.

Five classifiers were evaluated on spectral feature sets under LOCO cross-validation: logistic regression (LR), support vector machine with radial basis function kernel (SVM-RBF), gradient-boosted trees (XGBoost), k-nearest neighbors (KNN), and Gaussian naïve Bayes (GNB).

Classifier hyperparameters are fixed at:

| Model | Hyperparameters |
| --- | --- |
| Logistic Regression | L2, lbfgs, C=1, balanced class weight |
| Support Vector Machine | RBF kernel, C=1, balanced class weight |
| XGBoost | balanced class weight |
| K-Nearest Neighbours | k=5, distance-weighted |
| Gaussian Naive Bayes | var_smoothing=1e-9 |
