## Supplementary Material 2: Developing a benchmark for machine learning based localization of putative epileptogenic zones from human interictal intracr for "Localizing epileptogenic zones using interictal intracranial electroencephalography and deep learning"

**Supplementary Material 2: Developing a benchmark for machine learning based localization of putative epileptogenic zones from human interictal intracranial electroencephalography.**

Introduction

Accurate localization of the epileptogenic zone (EZ) is vital in the presurgical evaluation of drug-resistant focal epilepsy. Published machine learning (ML) models vary substantially in data source, electrode modality, feature engineering, and validation strategy. This makes their reported performance difficult to compare directly. As a result, there is a need for a literature-level benchmark that summarizes how well current approaches localize putative epileptogenic tissue from human interictal iEEG.

To address this gap, we systematically reviewed studies applying ML to human interictal iEEG for putative EZ or seizure onset zone localization, extracted performance metrics, and synthesized AUROC estimates using random-effects meta-analysis. We also examined performance by algorithm class, separating classical ML from deep learning, to place current approaches in a common quantitative frame of reference.

Method

A systematic search of PubMed was conducted up to May 2026 using a structured Boolean query combining MeSH headings and free-text terms “Epilepsy, Focal” [Mesh], “epileptogenic zone”, “seizure onset zone”, "EZ", “SOZ”, terms for intracranial electroencephalography modalities (sEEG, ECoG, subdural grids, depth electrodes), and machine learning methodology (“Machine Learning” [Mesh], “Artificial Intelligence” [Mesh], deep learning, neural networks, statistical machine learning, classical machine learning).

Studies were included if they met all of the following criteria: (1) human clinical data from patients with drug-resistant focal epilepsy undergoing presurgical evaluation with intracranial electrodes (SEEG or ECOG); (2) ML applied exclusively to interictal iEEG raw data or extracted features such as high-frequency oscillations, band power, entropy; (3) localization performance evaluated against clinical labels such as clinical neurophysiological annotation and post-operative seizure freedom; and (4) reporting of at least one quantitative discrimination metric with an accompanying sample size sufficient to derive variance. Studies were excluded if they used animal models, in silico data, scalp EEG recordings, ictal features, purely anatomical neuroimaging, or if they were reviews, letters, abstracts. Multiple distinct algorithmic conditions or patient cohorts reported within a single publication were treated as separate effect sizes, each attributed to the parent study with a disambiguating suffix (e.g., Klimes 2019a–j).

For each eligible study arm, the following data were extracted: first author, publication year, sample size, dataset origin, data recording modality, sampling rate, and duration of analyzed iEEG data. Clinical variables included epilepsy etiology, positive class nomenclature, and positive class definition. ML–related variables included the implemented algorithm, derived feature sets, dimensionality reduction techniques, feature selection methods, model evaluation approaches, and performance metrics, including sensitivity, specificity, accuracy, precision, F1-score, area under the receiver operating characteristic curve (AUROC), and area under the precision–recall curve (AUPRC). Values reported as “not reported” were coded as missing and excluded from the relevant analyses.

Random-effects meta-analysis of AUROC is implemented to derive a literature benchmark. Because AUROC is a bounded probability metric (range 0 to 1), each observed value $y_{i}$ was stabilized via the logit (log-odds) transformation prior to modelling:

$$ŷ_{i} = logit(y_{i} ) = log[\frac{y_{i}}{1-y_{i}}]$$

Within-study sampling variance for each logit-transformed estimate was approximated using the delta method:

$${\sigma^{2}}_{i} \approx\frac{1}{[n_{i} \cdot p_{i}\cdot(1 - p_{i})]}$$

where $n_{i}$ is the study-arm sample size and $p_{i}=\mathrm{expit}(\hat{y}_{i})=\frac{1}{1+e^{-\hat{y}_{i}}}$ is the back-transformed AUROC value. This derives study-specific, precision-weighted variance estimates.

Pooling was performed using the frequentist random-effects estimator in Python via statsmodels 0.14.6. The $I^{2}$ statistic computed to show the proportion of total variance attributable to between-study differences. All logit-scale estimates were back-transformed to the probability scale via the expit function to yield the pooled AUROC and its intervals.

To examine whether algorithm class moderated AUROC, the random-effects estimator was applied independently within each algorithm stratum classical ML (CML) and deep learning (DL) as separate subgroups. Stratum-specific pooled AUROC values and 95% CIs are reported. Because the subgroup analyses were performed as independent, within-stratum models rather than a single joint model, no formal cross-stratum statistical test was computed; instead, the degree of overlap between the 95% CIs of CML and DL is used as a descriptive indicator of between-class comparability. This approach mirrors standard practice for exploratory algorithm-class moderator analyses in small meta-analytic datasets where formal interaction tests are underpowered.

Results

24 studies implementing ML for identification of putative EZs from human interictal iEEG are identified. (Table 1, Table 2) Of which, 11 studies reported AUROC metric and are pooled by the random-effects model. Because individual papers frequently reported performance across multiple distinct algorithmic conditions or patient cohorts, the final meta-analytic dataset comprised 46 independent study arms (effect sizes).

The random-effects meta-analysis of 46 study arms yielded a pooled AUROC of 0.765 (95% CI: 0.743–0.787) on the back-transformed probability scale. (Figure 1) This estimate represents the expected discriminative performance of a ML model localizing the EZ from interictal iEEG and constitutes the benchmark value derived from the current literature.

Moderate between-study heterogeneity was detected with an approximate $I^{2}$ of 0.33. Approximately one-third of the total variability across study arms therefore reflects genuine between-study differences rather than within-study sampling error.

Independent random-effects meta-analyses within each algorithm stratum yielded a pooled AUROC of 0.758 (95% CI: 0.731–0.782) for CML (k = 42 arms) and 0.810 (95% CI: 0.694–0.889) for DL (k = 4 arms). Although DL approaches produced a numerically higher point estimate, the 95% CIs overlapped substantially, and the DL interval was considerably wider, reflecting the small number of DL arms currently available in the literature. The imprecision of the DL estimate precludes any definitive conclusion of superiority over CML. Therefore, both algorithm classes achieve broadly comparable discriminative performance, with a pooled benchmark AUROC in the range of approximately 0.76–0.81 across algorithms.

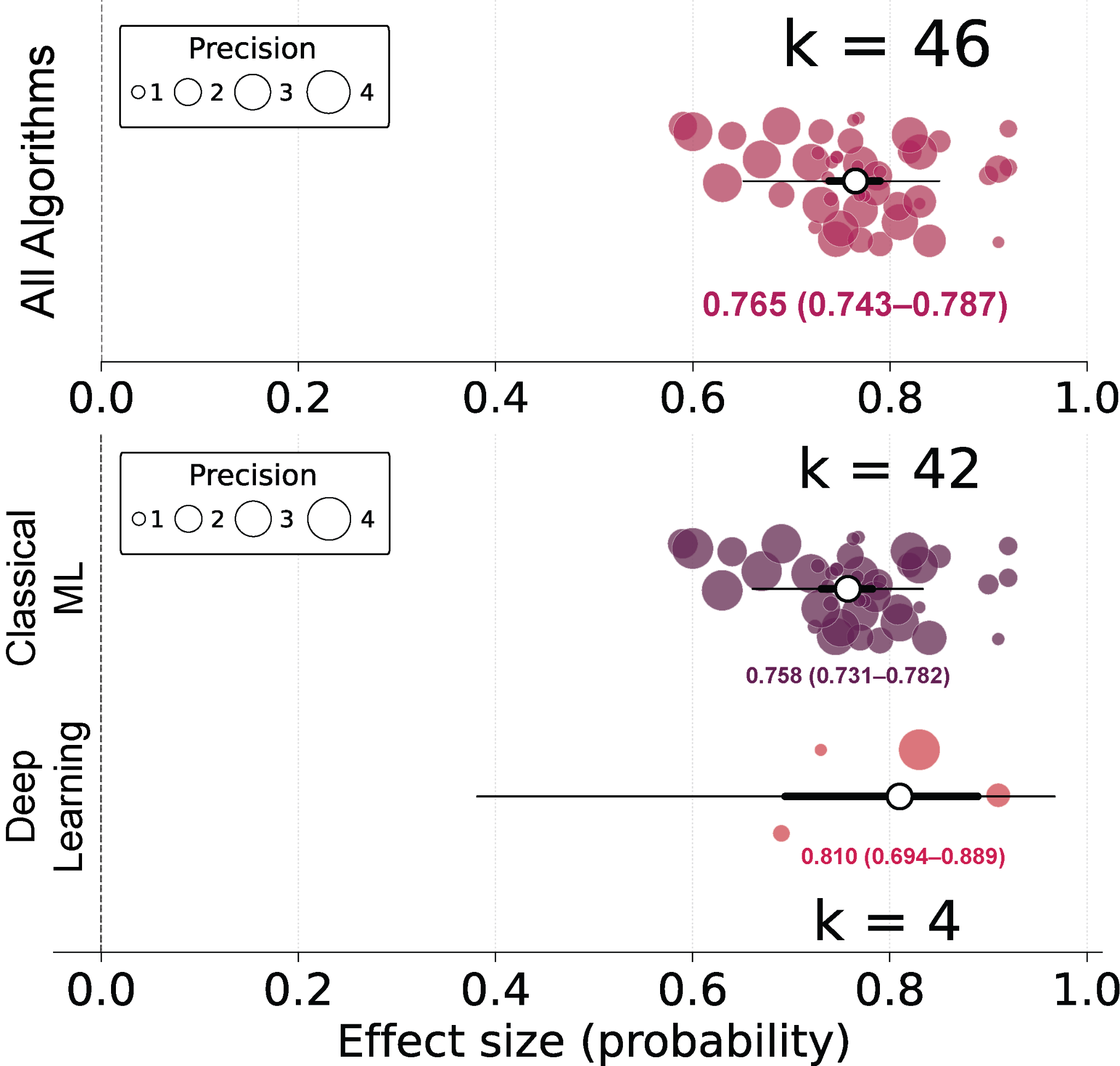

Figure 1. Orchard plot showing pooled mean and 95% CI estimates of AUROC

| Author Year | Data Publicity | Dataset | N | Age | Gender | Epilepsy Aetiology | Nomenclature | Definition | Seizure Free Only? | Follow up length | Recording State |
| --- | --- | --- | --- | --- | --- | --- | --- | --- | --- | --- | --- |
| Jose 2023 (1) | Private | Amrita Advanced Centre for Epilepsy | 15 | Range 11-43 | Not reported | Focal cortical dysplasia Reactive gliosis Dysplasia Hippocampal sclerosis  Non-diagnostic | Resected Zone | The region that was excised, resulting in seizure freedom | Yes | 44.4 months | NREM sleep |
| Cimbalnik 2019 (2) | Private | Brno Epilepsy Centre, St. Anne’s University Hospital  Mayo Clinic | 77 | Not reported | Not reported | Not reported | Seizure onset zone | Visual identification of a clear electrographic seizure discharge, followed by a look back in the iEEG recordings for the earliest electroencephalographic change contiguously associated with the seizure. | No | Not reported | Brno: awake Mayo: asleep |
| Chybowski 2024 (3) | Private | Montreal Neurological Institute and Hospital Brno Epilepsy Centre, St. Anne’s University | 25 | Median 28 IQR 13.5 | 14M 11F | Focal cortical dysplasia Gliosis | Epileptogenic zone | Resected seizure onset contacts in patients with good post-surgical outcome | Yes | >12 months | Wake, NREM sleep stages N2 and N3, REM sleep |
| Guo 2025 (4) | Both | OpenNeuro HUP iEEG Epilepsy Dataset  Xuanwu Hospital, China | 27 | Not reported | 13M 24F | Not reported | Seizure onset zone | Epileptologist determination | No | Not reported | Not reported |
| Makaram 2023 (5) | Private | Boston Children's Hospital Epilepsy Centre | 20 | Mean 12  Std 6 | 14M 6F | Focal cortical dysplasia  Gliosis  Unknown aetiology | Epileptogenic zone | Area of the brain that was resected during surgery | Yes | >12 months | Not reported |
| Xiao 2021 (6) | Private | Shenzhen Second People's Hospital | 3 | Mean 14.3  Std 3.68 | 2M 1F | Not reported | Seizure onset zone | Region of cortex from which seizures originate  Epilepsy specialist determination | Yes | >12 months | Not reported |
| Wang 2022 (7) | Private | Xuanwu Hospital, China | 10 | Range 20-38 | 4M 6F | Hippocampal sclerosis | Seizure onset zone | The first unambiguous visual signal alterations after seizure onset | Yes | Mean 16.4  Range 9-33 months | Asleep |
| Sumsky 2019 (8) | Public | National Institutes of Neurological Disease and Stroke iEEG Portal | 14 | Range 9-39 | 10M 4F | Cryptogenic  Meningitis  Traumatic brain injury  Dysplasia | Seizure onset zone | Resected volume in patients with successful outcomes, or clinically determined seizure onset zone if resected volume not available | No | 12 months | Both |
| Klimes 2019 (9) | Private | Montreal Neurological Institute and Hospital | 30 | Mean 34.2  Std 10.1 | 16M 14F | Malformation of cortical development Hippocampal sclerosis  Gliosis Tumour | Epileptogenic zone | Resected seizure onset zone contacts from good outcome patients | No | Mean 35.5  Std 14.9 months | Wake, NREM sleep stages N2 and N3, REM sleep |
| Conrad 2022 (10) | Public | ieeg.org | 101 | Median 36 Range 16-69 | 47M 54F | Lesional  Non-lesional | Seizure onset zone | Epileptologist determination | No | Not reported | Both |
| Klimes 2024 (11) | Private | Montreal Neurological Institute and Hospital  Brno Epilepsy Centre, St. Anne’s University | 50 | Median 32.5 IQR 11 | 27M 23F | Focal cortical dysplasia Gliosis | Epileptogenic zone | Resected area in patients with good surgical outcome | No | >12 months | NREM sleep |
| Chen 2015 (12) | Public | Bern-Barcelona | 5 | Not reported | Not reported | Not reported | Focal EEG channels | Channels that first show ictal EEG signal changes on visual examination | Not reported | Not reported | Not reported |
| Lundstrom 2021 (13) | Public | Mayo Clinic | 83 | Median 40  Range 5-75 | 30M 53F | Not reported | Seizure onset zone | Earliest EEG change in a clear electrographic seizure, determined by epileptologist | No | Median 5.4 Range 0.3-14.5 years | Asleep |
| Varotto 2021 (14) | Private | Claudio Munari Epilepsy Surgery Centre of Niguarda Hospital | 10 | Mean 31.7  Std 7.3 | 7M 3F | Cryptogenic Non-diagnostic Focal cortical dysplasia | Epileptogenic zone | Intersection between SEEG leads labelled as epileptogenic zone through pre-surgical evaluation and the resected zone | Yes | Mean 56  Std 13 months | Awake |
| Matsubayashi 2023 (15) | Private | Juntendo University Hospital | 11 | Range 5-39 | 7M 4F | Focal cortical dysplasia | Seizure onset zone | Epileptologist determination | No | Not reported | Not reported |
| Wang 2024 (16) | Private | Niguarda "Ca' Granda" Hospital, Milan, Italy | 65 | Mean 29.7  Std 9.5 | 35M 29F | Not reported | Seizure zone | Not explicitly stated | Not reported | Not reported | Training: awake  Test: asleep |
| Zhao 2022 (17) | Public | Bern-Barcelona Juntendo University Hospital | Not reported | Not reported | Not reported | Focal cortical dysplasia | Seizure onset zone | Epileptologist determination | Not reported | Not reported | Not reported |
| Miao 2022 (18) | Private | Juntendo University Hospital | 7 | Range 5-39 | 5M 2F | Focal cortical dysplasia | Seizure onset zone | Epileptologist determination | Yes | Mean 4  Std 0.886 months | Not reported |
| Akter 2020 (19) | Private | Juntendo University Hospital | 8 | Not reported | Not reported | Focal cortical dysplasia | Seizure onset zone | Epileptologist determination | Not reported | Not reported | Not reported |
| Varatharajah 2018 (20) | Private | Mayo Clinic | 82 | 31 | 48M 34F | Not reported | Seizure onset zone | Electrodes with the earliest iEEG seizure discharges on visual examination | No | Not reported | Asleep |
| Zhao 2022 (21) | Private | Juntendo University Hospital | 6 | Mean 22.7  Std 11.1 | 4M 2F | Focal cortical dysplasia | Seizure onset zone | Epileptologist determination | No | Mean 4.25  Std 0.901 months | Asleep |
| Nejedly 2025 (22) | Private | St. Anne's University Hospital in Brno | 80 | Mean 33  Std 10 | 46M 34F | Focal cortical dysplasia, hippocampal, gliosis, nodular heterotopia, various | Intervention zone | Contacts within the bounds of the surgical resection or radiofrequency thermocoagulation ablation | No | 1 year | Awake |
| Sundrani 2025 (23) | Private | Vanderbilt University Medical Center | 78 | Mean 31.4  Std 8.9 | 33M 45F | Various seizure types: FIAS, FAS, FBTC, GTC | Seizure onset zone | Epileptologist determination | No | 1 year | Awake |
| Pilet 2025 (24) | Private | Medical College of Wisconsin | 26 | Mean 35.5  Std 12.1 | 14M 12F | Malformations of cortical development, mesial temporal sclerosis, low-grade gliomas, vascular malformations, encephalomalacia | Seizure onset zone and resection zone | SOZ defined by epileptologist determination. The RZ was defined as electrodes within the resection and those within a 5 mm margin | Yes | Mean 7  Std 3.1 years | Awake |

Table 1. Characteristics of datasets used for development of current machine learning models in classifying putative epileptogenic zone channels.

| Author Year | Data modality | Sampling rate | Duration of analysed segments | Preprocessing steps | Features considered | Feature selection method | Supervised vs Unsupervised | ML algorithms | Class distribution | Class imbalance handling | Train-test split method |
| --- | --- | --- | --- | --- | --- | --- | --- | --- | --- | --- | --- |
| Jose 2023 (1) | SEEG | 1024, 2048 Hz | 1s window around interictal spikes | Bipolar montage  Bandpass: 0.5-70 Hz  Notch filter: 50 Hz | Amplitude, phase amplitude coupling, root mean square, power spectral density, Lyapunov exponent, correlation dimension, approximate entropy, in-degree, out-degree, and total degree | Not reported | Supervised | Support Vector Machines, Decision Trees, K Nearest Neighbours, Linear Regression, Random Forests, Gaussian Naive Bayes, Bagging Classifiers, Extra Trees, Ada Boosting, Gradient Boosting | Not reported | Not reported | 10-fold cross-validation |
| Cimbalnik 2019 (2) | iEEG | Brno: 25 kHz  Mayo: 32 kHz | Not reported | Downsample to 5kHz  Artifact removal: visual inspection, automated detection  Referencing | HFO rate, band power, frequency-amplitude coupling, phase-amplitude coupling, power spectral entropy, correlation, correlation with lag, correlation delay, phase synchrony, phase consistency, phase lag index, phase lag index delay, relative entropy | ANOVA with F-score values to determine relevant features | Supervised | Support Vector Machine | Not reported | Inverse proportional class weighting | Leave-One-Out Cross-Validation |
| Chybowski 2024 (3) | SEEG | MNI: 2000 Hz SAUH: 5000 Hz | 5mins | Average referencing | Spectral power, power spectral entropy, phase-amplitude coupling, frequency-amplitude coupling, low-frequency ratio, Hjorth complexity, phase synchrony, phase consistency, phase lag index, linear correlation, coherence, relative entropy, IED rates, HFO rates | Recursive Feature Elimination with Cross-Validation | Supervised | Logistic Regression, Support Vector Regression, ElasticNet | 15.2:1 | Not reported | Leave-One-Out Cross-Validation |
| Guo 2025 (4) | SEEG | 1024, 2048 Hz | 1s | Notch filtering  Low pass: 100 Hz  Re-referencing: zero mean  Rescaling: unit variance | Attribute vectors extracted by temporal convolutional network from SEEG channels, adjacency matrices representing functional connectivity between channels | Not applicable (deep learning) | Supervised | Adaptive Spatial-Temporal Graph Neural Network with graph convolutional network and long short-term memory | Balanced dataset created for each patient | Not applicable | One seizure for testing, others for training |
| Makaram 2023 (5) | iEEG | Not reported | 2mins | Bandpass: 1-70 Hz | Time-frequency images created using Morlet wavelet transform, followed by extraction of unsupervised activation energy (visual complexity) from 13 convolutional layers of a pre-trained VGG16 network | Not applicable (deep learning) | Supervised | Support Vector Machine with linear kernel | 1.42:1 | Not reported | Three-fold cross-validation |
| Xiao 2021 (6) | SEEG | 2048 Hz | 1s | Removal of interferences (MR gradient, eye movement pseudo difference)  Bandpass: 60-140 Hz | Normalised high-frequency energy extracted with finite impulse response filter | Not reported | Supervised | Convolutional Neural Network (seizure detection)  K-means clustering (SOZ localisation) | Not reported | Not reported | Not reported |
| Wang 2022 (7) | SEEG | 2048 Hz | 30 mins | Not reported | 126 features including spike rate, normalised pathological ripple rate, fast ripple rate, ripples co-occurring with fast ripples rate, and various energy features from time, frequency, and wavelet domains | Shapley value and hypothetical testing | Supervised | Deep Neural Network with attention mechanism | 2.91:1 | Cross entropy with focal loss training | Leave-One-Out Cross-Validation |
| Sumsky 2019 (8) | iEEG | 512 Hz | 10 mins | High pass: 80 Hz  Common average reference subtraction  Artifact removal | Ripple event rates (80-250 Hz) and rank-based susceptibility index | Not applicable | Supervised | Support Vector Machine with radial basis kernel | Imbalanced (distribution not reported) | Not reported | K-fold cross-validation with patient-level separation |
| Klimes 2019 (9) | SEEG | 2000 Hz | 30s | Common average reference subtraction  Bandpass filtering in eight frequency bands | Oscillatory events and HFO rates, power spectral entropy, power in band, linear correlation, linear correlation with lag, relative entropy, phase synchrony, phase consistency, phase-lag index, spike features | ANOVA with F-score, outliers determined using modified z-score with threshold set to 3 | Supervised | Support Vector Machine | Not reported | Class weighting | Leave-One-Out Cross-Validation |
| Conrad 2022 (10) | 18 ECoG 83 SEEG | Not reported | 1min | Spike detection | Spike rates in different brain states (wake, sleep, pre-ictal, post-ictal) | Not applicable | Supervised | Mixed-effects logistic regression | Not reported | Not reported | Leave-One-Out Cross-Validation |
| Klimes 2024 (11) | SEEG | 2000, 5000 Hz | 5mins | Average referencing | IED rates, gamma-IEDs, HFO rates, power in band, power spectral entropy, relative entropy, phase-amplitude coupling, frequency-amplitude coupling, low-frequency ratio, Hjorth complexity, phase synchrony, phase consistency, phase lag index, linear correlation, correlation with lag, correlation delay | Hierarchical clustering on Spearman rank-order correlations followed by ANOVA F-score values | Supervised | Logistic regression, SVM with linear kernel, SVM with RBF kernel, random forests | 10.2:1 | Inverse proportional class weighting | Leave-One-Out Cross-Validation |
| Chen 2015 (12) | Intracranial EEG (iEEG) | 512 Hz | 20s | Bandpass: 0.5-150 Hz | Maximum coefficient, minimum coefficient, mean of coefficients, standard deviation of coefficients, skewness of coefficients, kurtosis of coefficients, squared sum of all coefficients, normalised standard deviation, normalised energy | Exhaustive search of all feature | Supervised | Support Vector Machine with RBF kernel | Balanced | Not applicable | Leave-One-Out Cross-Validation |
| Lundstrom 2021 (13) | iEEG | 32 kHz |  | Downsample to 5kHz  Low pass: 1 kHz  Decimation to 250 Hz for low frequency analysis | 20-50 Hz to 0.02-0.5 Hz power ratio, interictal epileptiform discharge rates, HFO rates | Not applicable | Supervised | Logistic regression | Not reported | Not reported | Leave-One-Out Cross-Validation |
| Varotto 2021 (14) | SEEG | 1000 Hz | 5s | Bipolar montage  Artifact removal | Graph theory-based centrality measures: outdegree centrality, indegree centrality, outstrength centrality, instrength centrality, betweenness centrality, outcloseness centrality, incloseness centrality, pagerank centrality, and eigenvector centrality | Not reported | Supervised | Decision Tree, Discriminant Analysis, Logistic Regression, Naïve Bayes, Support Vector Machine, K-Nearest Neighbours, Boosted Ensemble, Bagged Ensemble, Discriminant Analysis Ensemble, KNN Ensemble | 7.87:1 | Oversampling: ADASYN, Adjusting the direction of the synthetic minority class example ADOMS, Random oversampling, Selective Pre-processing for Imbalanced Data, Borderline-Synthetic Monitoring Oversampling Technique   Underdamping: Condensed Nearest Neighbour, Neighbourhood Cleaning Rule, One Side Selection, Random Under sampling, Under sampling based on clustering | Leave-One-Out Cross-Validation |
| Matsubayashi 2023 (15) | iEEG | 2000, 1000 Hz | 20s | Not explicitly reported | Not detailed. 12 features extracted from EEG sub-bands. | Not applicable | Supervised | Domain adversarial neural networks, adversarial discriminative domain adaptation, maximum classifier discrepancy | Imbalanced (ratio not reported) | Not reported | Leave-One-Out Cross-Validation |
| Wang 2024 (16) | SEEG | Not reported | 10mins | Not explicitly detailed | 260 features: 60 criticality features (DFA exponent, functional E/I index, bistability index) across 20 frequency bands; 200 synchrony features: effective weight, eigenvector centrality, clustering coefficient, local efficiency across 50 frequency bands | Singular value decomposition for dimensionality reduction to rank-10 eigen-feature space | Unsupervised | Gaussian Mixture Models, Fuzzy C-means | Balanced | Not reported | Training on resting-state SEEG, testing on sleep SEEG |
| Zhao 2022 (17) | iEEG | Bern-Barcelona: 512 Hz Juntendo: 2000 Hz | 20s | Median re-referencing  Bandpass: 0.5-150 Hz (Bern-Barcelona) | 8 entropy features across 7 frequency bands, Short-time Fourier transform Time-frequency representations | Not reported | Semi-supervised learning | Support Vector Machine, Fully Connected Neural Network, Convolutional Neural Network, Positive-Unlabelled learning | Imbalanced (distribution not reported) | Random channel selection | 10-fold cross-validation  (within patient validation) |
| Miao 2022 (18) | ECoG | 2000 Hz | 20s | Bandpass: 0.5-24 Hz  High-frequency filter: 80-560 Hz with 30 Hz bandwidth | Phase-amplitude coupling values between low-frequency phase (0.5-24 Haz) and high-frequency amplitude (80-560 Hz) presented as comodulograms | Not applicable | Supervised learning | LightGBM, 2D CNN, SVM Linear, SVM RBF | 6.27:1 | CNN with weighted focal loss training SVM/LightGBM: Class weighting | Time series nest cross-validation |
| Akter 2020 (19) | ECoG | 2000 Hz | 20s | Bandpass: 100-600 Hz | Eight entropy measures: Approximate Entropy, Permutation Entropy, Shannon Entropy, Sample Entropy, Tsallis Entropy, Phase Entropy 2, Phase Entropy 1, and Renyi's Entropy | Sparse Linear Discriminant Analysis | Supervised | Support Vector Machine with radial basis function kernel | Not reported | Adaptive Synthetic Sampling Approach | 10-fold cross-validation |
| Varatharajah 2018 (20) | SEEG | 5000 Hz | 3s epoch accommodating at least a single transient electrophysiologic event (PAC, HFO, IED) | Notch filter: 60 Hz  Artifact removal | Local rates of high frequency oscillations, interictal epileptiform discharges, and phase amplitude coupling within 10-minute windows | Clustering procedure to identify normal/abnormal channels based on biomarker measures | Supervised | Support Vector Machine with RBF kernel | 4.45:1 | Not reported | Leave-One-Out Cross-Validation |
| Zhao 2022 (21) | ECoG | 2000 Hz | 10s | Bandpass: 0.5-900 Hz | Representation learning using deep learning | Not applicable (deep learning) | Supervised | One-dimensional Convolutional Neural Network | 5.73:1 | Data augmentation: discrete cosine transforms, ADASYN, SMOTE Class-balanced focal loss | Leave-One-Out Cross-Validation |
| Nejedly 2025 (22) | SEEG | 5000 Hz | 30 minutes continuous | Lowpass filtering | Univariate features were extracted from the SEEG recordings using a custom pipeline. Functional connectivity was derived from a spike propagation matrix. Euclidean distances between contacts were calculated to create graph edges. | Not applicable (deep learning) | Supervised | Graph Attention Network | Imbalanced (distribution not reported) | Not reported | Leave-One-Out Cross-Validation |
| Sundrani 2025 (23) | SEEG | Not reported | 30s | Bandpassing: 1–59 Hz, 61–119 Hz, and 121–150 Hz.  Input windows were normalized via interquartile range (IQR)/median normalization. | Feature-agnostic end-to-end deep learning. | Not applicable (deep learning) | Supervised | One-dimensional Convolutional Neural Network | Imbalanced (distribution not reported) | Not reported | 5-fold-Cross-Validation |
| Pilet 2025 (24) | iEEG | 1000 Hz | 12s | Visual rejection of channels with excessive noise; removal of power line noise (60 Hz) and its first five harmonics via spectrum estimation; re-referencing to the common average reference; and bandpass filtering (two-pass, zero phase FIR filter). | Spike detection; HFO detection; power spectral density; functional connectivity (amplitude envelope correlation and phase-locking value using the Hilbert transform; followed by graph metric derivation (node strength, eigencentrality). | Feature subsets were grouped for targeted comparisons (e.g., gamma band vs. low frequency, AEC vs. PLV, or graph measures vs. PSD/Spike/HFOs) | Supervised | Support vector machine with a gaussian kernel. | Imbalanced (distribution not reported) | Not reported | Node-level 4-fold CV, 4-fold-Cross-Validation, Leave-One-Out Cross-Validation |

Table 2. Computational pipelines adopted by current machine learning studies.
