## Supplementary Material 3: Model hyperparameters for "Localizing epileptogenic zones using interictal intracranial electroencephalography and deep learning"

Temporal encoder

| Hyperparameter | Definition | Value used |
| --- | --- | --- |
| Number of wavelet frequency bands | How many frequency compositions the pre-processed signal is decomposed into before representation learning occurs | 6 |
| Wavelet cycles | Controls the time–frequency trade-off of each Morlet wavelet | 5 |
| Epoch length | The recording is divided into non-overlapping windows of this duration; each window is encoded independently before being aggregated | 10 |
| Temporal tokens per epoch | Each 10-second epoch is summarized into this many time-points before entering the within-epoch transformer | 8 |
| Embedding size | The width of internal representation | 64 |
| Dropout (temporal) | Randomly zeroes a fraction of activations during training to prevent the model memorizing the training data | 0.2 |
| Within-epoch transformer layers | Depth of the small transformer that processes each 10-second epoch | 1 |
| Within-epoch attention heads | Number of parallel attention mechanisms inside each transformer layer | 1 |
| Aggregator attention heads | Number of parallel attention mechanisms used when the model decides which 10-second epochs are most important for classifying a channel | 2 |
| Aggregator dropout | Regularization applied within the cross-epoch attention step | 0.1 |

Spatial encoder

| Hyperparameter | Definition | Value used |
| --- | --- | --- |
| Number of spatial layers | How many rounds of cross-channel communication are applied; each layer updates channel representation based on others temporal embedding | 2 |
| Spatial attention heads | Number of parallel attention mechanisms for spatial modelling | 2 |
| Number of inducing points | A bottleneck of learned surrogate channels that mediate communication between all real channels; avoids the computational cost of all-to-all attention | 32 |
| SwiGLU hidden dimension | Width of the feedforward network applied to each channel after attention, set to 8/3 emb_size following the SwiGLU convention | 171 |
| Spatial dropout | Regularization applied within the feedforward step of each spatial layer | 0.2 |
